# Monocyte chemoattractant protein-1 levels are associated with major depressive disorder

**DOI:** 10.1101/2020.11.26.20239293

**Authors:** Maliha Afrin Proma, Sohel Daria, Salsabil Islam, Zabun Nahar, Sardar Mohammad Ashraful Islam, Mohiuddin Ahmed Bhuiyan, Md. Rabiul Islam

**Author notes:** **Correspondence: Md. Rabiul Islam, PhD**, Assistant Professor, Department of Pharmacy, University of Asia Pacific, 74/A Green Road, Farmgate, Dhaka-1215, Bangladesh. These authors contributed equally (Maliha Afrin Proma, Sohel Daria).

## Abstract

Major depressive disorder (MDD) is a distressing condition characterized by persistent low mood, loss of interest in daily activities. Many biological, psycho-social, and genetic factors are thought to be involved with depression. The present study aimed to investigate the serum levels of monocyte chemoattractant protein-1 (MCP-1) in MDD patients to explore its role in the development of depression. This case-control study recruited 114 MDD patients and 106 healthy controls (HCs) matched by age and gender. A specialized psychiatrist diagnosed the cases and evaluated the controls based on the diagnostic and statistical manual for mental disorders, 5th edition. The serum MCP-1 levels were quantified by commercially available enzyme-linked immune sorbent assay kits. The Hamilton depression rating scale (Ham-D) was applied to measure the severity of depression. We observed the decreased levels of serum MCP-1 in MDD patients compared to HCs. A significant negative correlation was obtained between serum MCP-1 levels and Ham-D scores. Also, female MDD patients with higher Ham-D scores exhibited lower serum MCP-1 levels. The receiver operating characteristic analysis demonstrated the good diagnostic value of MCP-1 with the area under the curve at 0.837. The depression-related alteration of serum MCP-1 may be more complicated than the current assumption and depends on the characteristics of the individual patients. Our study suggests that the serum MCP-1 levels might be involved in the pathophysiology and mechanism of MDD. The present findings, along with the diagnostic evaluation, might be used to assess the depression risk.

## 1. Introduction

Major depressive disorder (MDD) is a chronic, highly prevalent, multi-symptomatic psychiatric mood disorder that affects around 350 million people around the world (WHO, 2017). Depression is all set to become the leading cause of disability worldwide suggested by epidemiological studies (WHO, 2017). Major depression alters a person’s perception and interaction with his surrounding environment, affecting the person’s personal as well as professional life (Zuckerman et al., 2018). The lifetime prevalence of MDD is about 15-20%, and currently, the diagnosis of MDD is performed based on clinical symptoms with a lack of biological markers (Li et al., 2018).

Several pathophysiological mechanisms have been postulated to understand this psychiatric disorder more clearly that include alterations in monoaminergic neurotransmission, imbalance of excitatory and inhibitory signaling in the brain, hyperactivity of the hypothalamic-pituitary-adrenal (HPA) axis, and changes in normal neurogenesis (Anjum et al., 2020; Emon et al., 2020; Islam et al., 2020; Islam et al., 2018a; Maes et al., 1993a; Varghese and Brown, 2001). Yet, the pathogenesis of MDD remains unclear because previous findings only led to a deeper understanding of the mental illness. Despite having various proposed mechanisms of developing MDD, mounting pieces of evidence have suggested a strong association between chronic inflammatory processes and the pathophysiology of MDD (Miller et al., 2009; Nishuty et al., 2019). Recent findings in neuroscience also indicated an association between the families of immune protein-designated chemokines and many neuroimmune processes related to psychiatric illnesses, for examples, synaptic transmission, and plasticity, hindrance in the progression of neurogenesis, and neuron-glia communication (Das et al., 2020; Heinisch and Kirby, 2009; Pujol et al., 2005; Stuart and Baune, 2014). Activation of the inflammatory response system causes alterations in any of these functions that may lead to the pathogenesis of MDD (Daria et al., 2020; Islam et al., 2018b; Nishuty et al., 2019).

Monocyte chemoattractant protein-1 (MCP-1) has attracted attention among other chemokines due to its immunomodulatory functions (Pae, 2014). MCP-1 is highly expressed in the neural areas of the brain region, for example, the cerebral cortex, hippocampus, and hypothalamus are important in the pathogenesis of MDD (Banisadr et al., 2005). The modulation of neural processes and neuroendocrine functions are commonly observed in MDD patients (Ali et al., 2020; Pae, 2014). MCP-1 is also known for its pleiotropic actions having functions in chemotaxis, stimulating functions on monocytes/macrophages, T lymphocytes, dendritic cells, and specific functions on the central nervous system (CNS) (Stuart and Baune, 2014). Besides, MCP-1 is found to be involved in the regulation of other cytokines that have been consistently reported as having an association in developing MDD (Pae, 2014). MCP-1 is produced by different types of cells including fibroblasts, astrocytes, monocytes, smooth muscle cells, endothelial cells, and microglial cells either constitutively or after stimulation by oxidative stress, cytokines, or by different growth factors (Deshmane et al., 2009). MCP-1 regulates the movement and permeation of monocytes, memory T lymphocytes, and natural killer cells through the regulation of both the T helper-1 (TH-1) and TH-2 systems (Sorensen et al., 2004; Traynor et al., 2002). By regulating the monocytes and TH systems, MCP-1 may alter both the innate and adaptive immune systems (Gu et al., 2000). It is plausible that MCP-1 is associated with the pathophysiology of MDD supporting the TH-1/TH-2 theory of developing depression (Schwarz et al., 2001). Psychomotor retardation is one of the clinical symptoms that is used in the diagnosis of depression (American Psychiatric Association, 2013). Inflammatory cytokine MCP-1 has a consistent association with psychomotor activity (Goldsmith et al., 2020). Goldsmith et al reported that the peripheral level of MCP-1 is correlated with the development of major depression (Goldsmith et al., 2016).

Although extensive researches were carried out to understand the neurobiological mechanisms of MDD, still the disease is being diagnosed based on the subjective symptoms and feedback of patients. Therefore, investigating biological markers might be helpful in the diagnosis and management of MDD. Taking all these together, the present study aimed to evaluate the peripheral levels of MCP-1 in MDD patients.

## 2. Methods

### 2.1 Participants

This case-control study recruited 114 MDD patients from the department of psychiatry of Bangabandhu Sheikh Mujib Medical University (BSMMU), Dhaka, Bangladesh. The present study protocol was approved by the respective institutional ethical review committee. The MDD cases were confirmed by a qualified psychiatrist following the diagnostic and statistical manual of mental disorders, fifth edition (DSM-5) (American Psychiatric Association, 2013). A total of 106 healthy controls (HCs) were enrolled from different parts of Dhaka city, Dhaka, Bangladesh, matched by age, gender, and body mass index (BMI) with MDD patients. The 17-item Hamilton depression rating scale (Ham-D) was applied to measure the severity of depression (Hamilton, 1960). HCs were also screened for any psychiatric disorders by interviewing with the same structured questionnaire that applied for MDD patients. The population with age range 18-60 years and having BMI within 16-34 kg/m2 were included in the present study. Exclusion criteria were mental retardation, presence of comorbid psychiatric illnesses, pregnancy, patients suffering from inflammatory diseases or taking any anti-inflammatory medication, diabetes, cardiovascular disorders, hypertension, excessive obesity, abnormal lipid profile, alcohol, and other substance abuse. A structured pre-designed questionnaire was used to record the socio-demographic profile of the study population. Every participant was well informed about the purpose and objective of the study and written consent was obtained from each of them before inclusion in the present investigation.

### 2.2 Blood sample collection

Five-milliliter of the blood sample was withdrawn from the cephalic vein of each participant by venipuncture and collected in a falcon tube. The collected samples were allowed to clot at room temperature for one hour. Then, the serum extraction was done by centrifugation of the samples at 1000 x g for 15 minutes. The serum was separated carefully from the centrifuged blood sample and stored at -80°C until further analysis.

### 2.3 Serum sample analysis

On the day of analysis, all the samples and reagents were equilibrated at room temperature. The serum samples were then vortexed for homogenous liquefaction. The enzyme-linked immunosorbent assay (ELISA) kit was used to measure the serum MCP-1 levels following the manufacturer’s instructions (Boster Biological Technology, USA). Briefly, 100µl of the diluted standards and samples were added into the appropriate wells of pre-coated plates. The plates were covered with the plate sealer and incubated for 90 minutes at 37 °C temperature. The cover was then removed and the liquid in the wells was discarded into an appropriate waste receptacle. Then, 100µl of prepared biotinylated anti-human MCP-1 antibody was added to each well. Again, the plates were covered with plate sealers and incubated for 60 minutes at 37°C temperature. Then, the liquid in the wells was discarded again and washed the plates 3 times with phosphate buffer. Then, 100µl of the prepared avidin-biotin-peroxidase complex was added to each well. After incubation of 40 minutes at room temperature, the liquids in the wells were again discarded and washed the plates 5 times with the wash buffer. At this stage, 90μl of color developing reagent was added to each well and incubated in the dark for 30 minutes at room temperature. Finally, 100µl of stop solution was added to each well and the absorbance was taken with a microplate reader at 450nm within 30 minutes. The concentration of serum MCP-1 was expressed as pg/mL. The assay was specific to natural and recombinant human MCP-1 with a sensitivity of less than 1pg/mL. To avoid heterogeneity in results, the analysis was carried out by the same persons and the serum levels of MCP-1 were measured simultaneously for both MDD patients and HCs.

### 2.4 Statistical analysis

The statistical package for the social sciences (SPSS) version 25.0 (Armonk, NY: IBM Corp.) was used for performing the necessary statistical analyses. The independent sample t-test and Fisher’s exact test were carried out for continuous variables and categorical variables, respectively. Spearman’s correlation test was used to find out the correlation between the analyzed parameter and severity score of depression. The box-plot graph was used to illustrate the variations of serum MCP-1 levels between the groups. The gender-specific scatterplot graph was applied to demonstrate the associations between serum MCP-1 levels and Ham-D scores in depression. Descriptive statistics were computed for the socio-demographic characteristics of the study population and data were expressed as mean ± standard error mean (SEM). Results were considered statistically significant when the p-value is less than or equal to 0.05.

## 3. Results

### 3.1 General description of the study population

The study population was categorized based on their socio-demographic profiles, biophysical characteristics, and smoking habit (Table 1). Both MDD patients and HCs were found similar in terms of socio-demographic profiles and biophysical characteristics. Females constitute the highest portion than males in both MDD patients and HCs. BMI values were normal for half of the study population. It was found that most of the MDD patients (87%) and HCs (92%) were literate. Most of the study population (60%) were from medium economic status. We observed most of the study population were non-smoker (MDD patients: 74%; HCs: 77%).

**Table 1.** Socio-demographic characteristics of the study population.

| Parameters | MDD patients (n=114) | Healthy controls (n=106) | *p value* |
| --- | --- | --- | --- |
| | Mean ± SEM | Mean ± SEM | |
| Age | 32.13 ± 0.84 | 33.88 ± 0.90 | 0.159 |
| 18-24 | 40 (35%) | 34 (32%) |  |
| 25-34 | 43 (38%) | 39 (37%) |  |
| 35-44 | 23 (20%) | 21 (20%) |  |
| 45-60 | 8 (7%) | 12 (11%) |  |
| Gender |  |  | 0.475 |
| Male | 54 (47%) | 47 (44%) |  |
| Female | 60 (53%) | 59 (56%) |  |
| BMI (kg/m<sup>2</sup>) | 25.20 ± 0.45 | 24.55 ± 0.37 | 0.280 |
| Below 18.5 (CED) | 6 (5%) | 3 (3%) |  |
| 18.5–25 (normal) | 57 (50%) | 59 (56%) |  |
| Above 25 (obese) | 51 (45%) | 42 (44%) |  |
| Monthly income (KBDT) | 67.97 ± 2.91 | 81.56 ± 4.11 | 0.195 |
| Below 30 | 5 (4%) | 27(25%) |  |
| 30–60 | 52 (46%) | 20(19%) |  |
| 61–90 | 36 (32%) | 27(25%) |  |
| Above 90 | 21 (18%) | 32(30%) |  |
| Occupation |  |  | 0.364 |
| Service | 15 (13%) | 11 (10%) |  |
| Business | 4 (4%) | 20 (19%) |  |
| Student | 3 (3%) | 23 (22%) |  |
| Jobless | 13 (11%) | 2(2%) |  |
| Others | 79 (69%) | 50 (47%) |  |
| Education level |  |  |  |
| Illiterate | 15 (13%) | 8 (8%) |  |
| Primary | 22 (19%) | 3 (3%) |  |
| Secondary | 53 (44%) | 43 (41%) |  |
| Graduate and above | 24 (21%) | 52 (49%) |  |
| Economic status |  |  | 0.235 |
| Low | 34 (30%) | 27 (25%) |  |
| Medium | 68 (60%) | 65 (61%) |  |
| High | 12 (11%) | 14 (13%) |  |
| Smoking habit |  |  | 0.585 |
| Yes | 30 (26%) | 24 (23%) |  |
| No | 84 (74%) | 82 (77%) |  |

BMI, body mass index; CED, chronic energy deficiency; KBDT, kilo Bangladeshi taka; MDD, major depressive disorder; SEM, standard error mean.

### 3.2 Comparison of serum MCP-1 levels in MDD patients and HCs

The clinical information and laboratory findings of the study population have been presented in Table 2. The serum concentrations of MCP-1 were 90.10±5.28pg/mL and 111.63 ±5.53 pg/mL for MDD patients and HCs, respectively, whereas the values of Ham-D score were 16.22 ± 0.46 and 4.63 ± 0.25MDD patients and HCs, respectively. We observed a statistically significant decrease in serum MCP-1levels in MDD patients compared to HCs (Fig. 1). Moreover, gender-wise serum MCP-1 levels among the study population showed a significant decrease in female MDD patients (91.69 ± 7.70 pg/mL) compare to female HCs (118.36 ± 8.73) but no such differences were seen in the male gender.

**Table 2.** Clinical information and laboratory findings of the study population.

| Parameters | MDD patients (n=114)<br>Mean ± SEM | Healthy controls (n=106)<br>Mean ± SEM | *p value* |
| --- | --- | --- | --- |
| Ham-D score | 16.22 ± 0.46 | 4.63 ± 0.25 | <0.001 |
| Serum MCP-1 level (pg/mL) | 90.10±5.28 | 111.63 ±5.53 | 0.005 |
| MCP-1 in female(pg/mL) | 91.69±7.70 | 118.36 ±8.73 | 0.024 |
| MCP-1 in male (pg/mL) | 88.67±7.31 | 106.26 ±7.09 | 0.087 |

Ham-D, 17-item Hamilton depression rating scale; MCP-1, monocyte Chemoattractant protein-1; MDD, major depressive disorder; SEM, standard error mean.

**Fig. 1.**
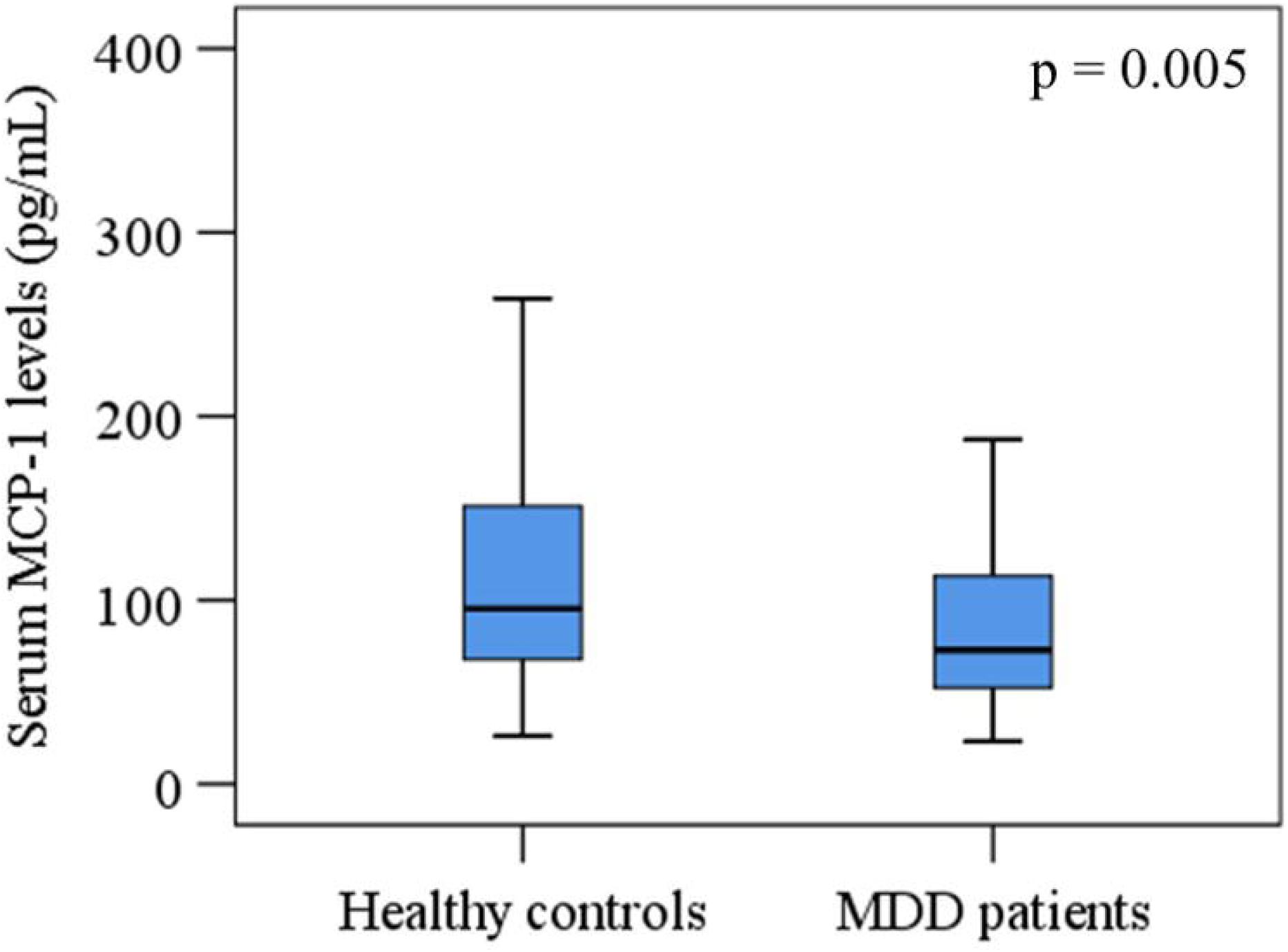
Variations of serum monocyte chemoattractant protein-1 (MCP-1) among the study population. A significant difference between the patient and control groups at a 95% confidence interval (CI).

### 3.3 Correlation between serum MCP-1 levels and Ham-D scores

Spearman’s correlation analysis was performed in the study population to find relationships among various research parameters. Serum MCP-1 levels were negatively correlated with the severity of depression (r = -479; p < 0.001). Moreover, females with higher Ham-D scores showed lower serum MCP-1 levels whereas males demonstrated the opposite results (Fig. 2). No such relationships were observed in serum MPC-1 levels or Ham-D scores with sociodemographic features.

**Fig. 2.**
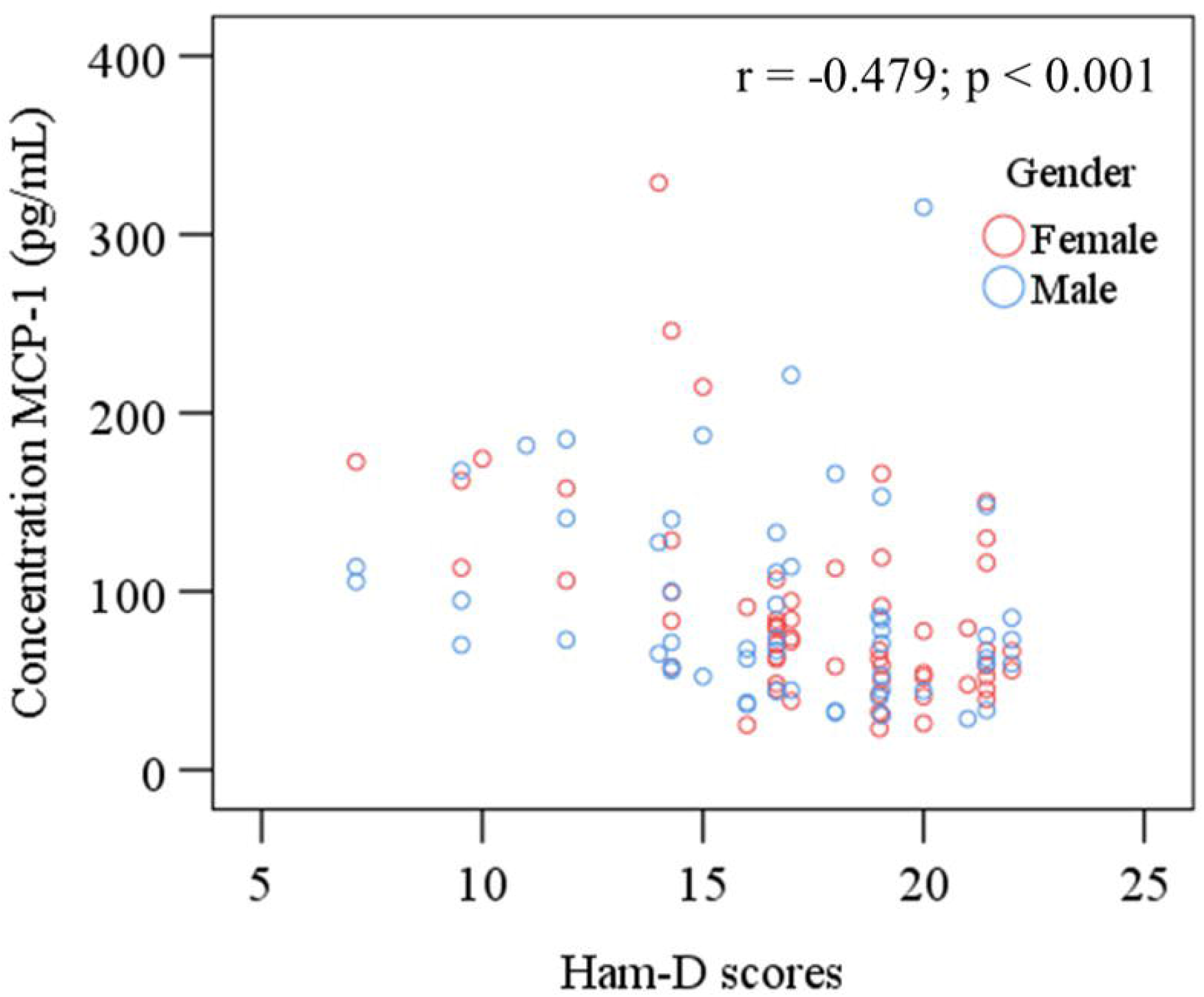
Scatter plot of serum monocyte chemoattractant protein-1 (MCP-1) and Ham-D scores with gender differences among MDD patients.

### 3.4 Evaluation of diagnostic value for serum MCP-1 levels

The ROC curve was plotted for serum MCP-1 levels and the cut-off point was detected for diagnostic purposes as 52.18 pg/mL (Fig. 3). The area under the ROC curve (AUC) was 0.837 which was significant (p < 0.05). The disease condition represents the lower value. According to the ROC analysis, the sensitivity, specificity, positive predictive value (PPV), and negative predictive value (NPV) were 80.1%, 82.5%, 79.4%, and 73.2%, respectively.

**Fig. 3.**
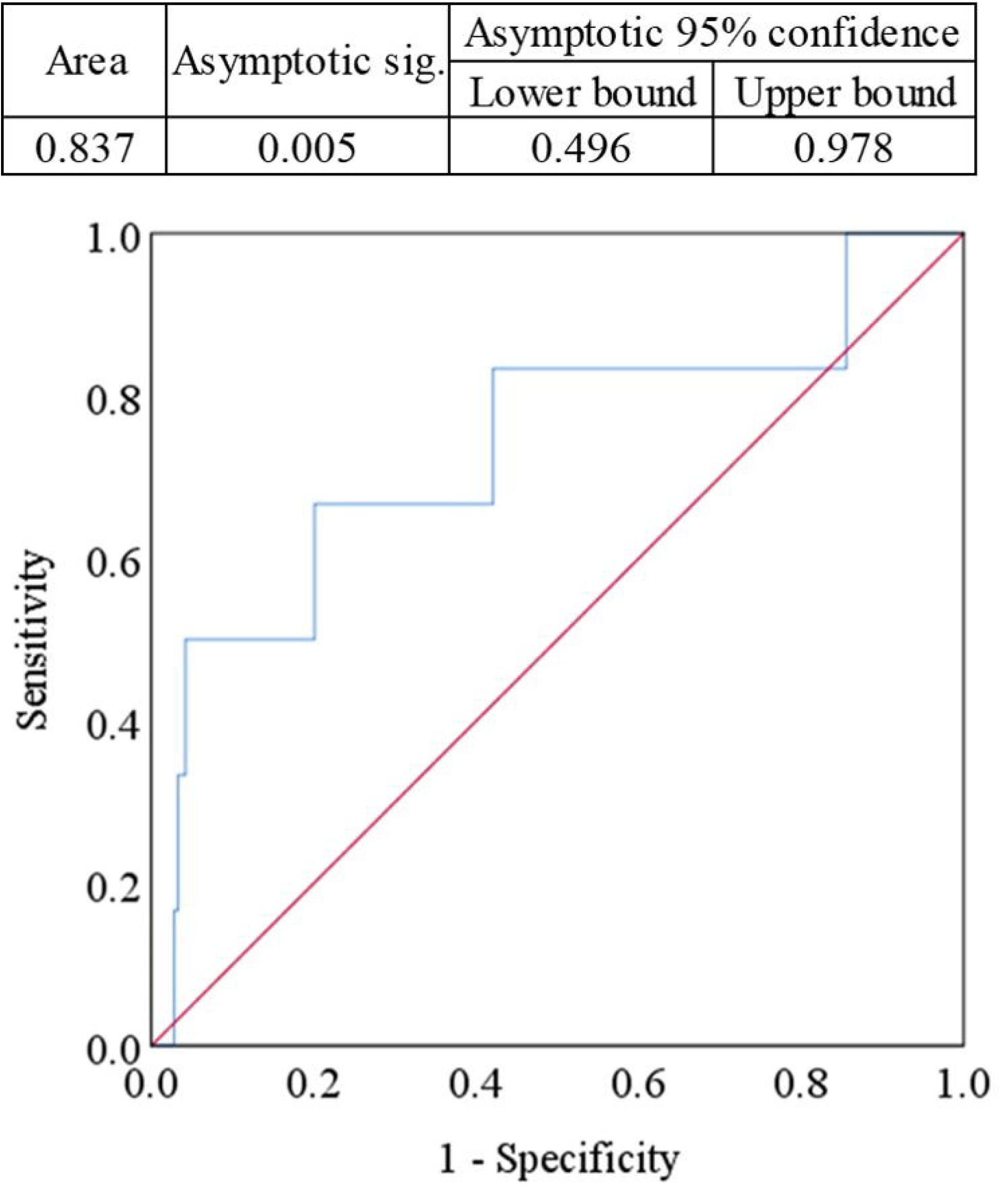
Receiver operating characteristic (ROC) curve for serum monocyte chemoattractant protein-1 (MCP-1). The cut-off point was detected as 52.18 pg/mL.

## 4. Discussion

To the best of our knowledge, this is the first-ever study among the Bangladeshi population attempted to determine the serum levels of MCP-1 in MDD patients to find the possible association between serum levels and pathophysiology of depression. The investigation of peripheral biological markers for mental disorders has been underway for many years (Domenici et al., 2010). Despite extensive investigations, a noninvasive blood-based test for the diagnosis of such diseases is still absent (Vetulani and Nalepa, 2000). The present study found significantly lowered serum MCP-1 levels in MDD patients compared to HCs. This alteration in the concentrations of serum MCP-1 might be activated by the inflammatory responses. Previous studies by Lehto et al and Grassi-Oliveria et al observed that reduced serum levels of MCP-1 in MDD patients compared to HCs were correlated with the severity and pathophysiology of depression (Grassi-Oliveira et al., 2012; Lehto et al., 2010). A recent investigation by Myungl et al found lower levels of chemokine MCP-1 in MDD patients compared to HCs (Myung et al., 2016). The actual mechanism behind these alterations and associations between MCP-1 levels and depression is still unclear. The possible explanations might be neuroprotective functions of the chemokines and their increased dopaminergic activity affects the central nervous system (CNS) (de Haas et al., 2007; Madrigal et al., 2009).

Neuroinflammation plays an important role in the pathophysiology of psychiatric disorders and alteration in the chemokine levels in MDD patients (Edberg et al., 2020). Chemokine MCP-1 performs functions in leukocyte trafficking, fibrosis, tissue remodeling, and angiogenesis during the neuroinflammation process (Lukacs, 2001; Schall and Bacon, 1994). MCP-1 binds with the C-C chemokine receptor type 2 (CCR2), which is present in different types of cells including astrocytes, microglia, neuronal stem cells, and neurons in CNS, and CCR2’s expression is stimulated in response to other pro-inflammatory cytokines (Andjelkovic et al., 2002; Banisadr et al., 2005; Banisadr et al., 2002; Croitoru-Lamoury et al., 2003; Guillemin et al., 2003). Macrophages, monocytes, dendritic cells, and T lymphocytes, all of which are involved in the inflammatory processes mainly activated by the chemokine MCP-1 (Deshmane et al., 2009). Some clinical trials investigated that the production of prostaglandin-E and proinflammatory cytokines were blocked by the antidepressant effect of COX-2 inhibitors (Abbasi et al., 2012; Akhondzadeh et al., 2009; Müller et al., 2006). Consequently, reduced levels of chemokines may have an association with the decreased functions of neurotransmitters in the CNS (Miller et al., 2009). Moreover, a preliminary study finding reported that the decreased concentrations of serum MCP-1 levels in MDD patients might be responsible for their suicidal ideation (Grassi-Oliveira et al., 2012).

Moreover, the current study performed the ROC curve analysis to quantify the predictive performance of altered MCP-1 levels in depression (Fig. 3). Area under the curve (AUC) in ROC analysis is used to determine the accuracy of prediction as follows: 0.90-1.0 = excellent, 0.80-0.90 = good, 0.70-0.80 = fair, 0.60-0.70 = poor and less than 0.6 = not useful (Won et al., 2018). The AUC was detected as 0.837 for the present investigation which was significant (p < 0.05). The lower values were assigned as the disease state and the cut of value was measured at 52.18 pg/mL. Many previous studies measured similar markers in depression but they did not evaluate the diagnostic performance of altered levels (Dalgard et al., 2017; Goldsmith et al., 2016). In this context, the present study is unique compared to past studies.

Additionally, several previous studies came up with the altered serum cytokine levels in depression but they could not suggest new therapeutic approaches based on their findings (Dahl et al., 2014; Gaarden et al., 2018). Approximately half of MDD patients do not respond to traditional antidepressant therapy (Bschor et al., 2012). This is happened due to the lack of efficacy or intolerable side effects of antidepressant molecules especially for those who have underlying immune deregulation (Rush et al., 2006; Trivedi et al., 2006). The high rate of treatment resistance may be responsible for the rising suicidal tendency of MDD patients and the unbearable economic burden to society (Pettorruso et al., 2020). Taken all these together, the present study proposed some new therapeutic approaches to improve the quality of life or even cure MDD patients. The approaches include pharmacotherapy, psychotherapy, and somatic therapy often employed for treatment and management of resistant depression which might manage the alteration of MCP-1 levels in MDD patients (Holtzheimer and Nemeroff, 2008). The repetitive transcranial magnetic stimulation (rTMS)and exercise are non-pharmacological synergistic approaches that can reduce stress, modulate immunity, improve the quality of life (QoL) of depressive patients (McGirr et al., 2015). Another relatively safe and effective option to manage depression can be vagus nerve stimulation (VNS) therapy (Zhang et al., 2020). Moreover, an innovative therapy could be immune activation where immunomodulatory effects help to restore the neurotransmitter synthesis and thus reduce the severity of depression (Ellul et al., 2020). Several activities related to neuropsychiatric disturbances might be exerted by MCP-1 including neuromodulator and neurotransmitter-like effects to regulate neurogenesis (Trettel et al., 2020). One of the studies found that MCP-1 is positively associated with cortical thickness in the anterior cingulate cortex (Poletti et al., 2019). Lehto et al. reported no difference in MCP-1 levels after treating MDD patients with antidepressants, anxiolytic, and antipsychotic medications when compared with other MDD patients (Lehto et al., 2010). But a recent study by Edberg et al. revealed that MCP-1 is associated with both pro-and anti-inflammatory analytes in depressive subjects. During cyclooxygenase-2 (COX-2) inhibition treatment, alteration in MCP-1 levels influenced by its surrounding inflammatory milieu (Edberg et al., 2020).

This is the first-ever study carried out in Bangladesh with such a large and homogeneous sample where age and gender were strictly matched for cases and controls. The same experimental conditions were maintained during the simultaneous measurement of MCP-1in MDD patients and HCs. The serum monocyte chemoattractant protein-1 in MDD patients compared to healthy controls aided in the field of biological psychiatry for the proper diagnosis and management of MDD. The present study didn’t consider the food habits, sleep pattern, current medication, nicotine users from the study population that may be potential confounding factors for chemokine MCP-1 analysis. Also, we did not evaluate the effects of antidepressant medications on serum MCP-1 levels.

The present study findings should be treated as preliminary and future studies with a large and more homogeneous population are required in a similar field to obtain better results. Research involving pre-and post-measurement of serum MCP-1 could be able to assess the medication effects on depression. Future research can be carried out involving the effects of mind-body therapies (MBTs) and physical exercises on peripheral MCP-1 levels in MDD patients to see the effectiveness of the suggested therapeutic approaches.

## 5. Conclusion

Our findings suggest that depression-related reduction of serum MCP-1 may be more complicated than previously presumed and could vary based on the characteristics of individual MDD patients. The present study demonstrated a possible association between reduced levels of MCP-1 and the pathophysiology of depression. The findings of the study along with their predictive performance could be used to determine depression risk and management of MDD.

## Data Availability

Data supporting the present study findings are within this article. All the relevant data and information can be obtained from the corresponding author upon reasonable request.

## Declarations

### Ethical consideration

The ethical review committee of the department of psychiatry, BSMMU, reviewed, and approved this study protocol (reference BSMMU/2019/3507). The principles mentioned in the Declaration of Helsinki were strictly followed during performing all the investigations.

## Acknowledgments

All the authors acknowledge the participants for their cooperation to conduct the study.

## Author’s contribution

MAP conceived the idea, designed the study, collected, and analyzed the data. SD interpreted the data, drafted, and revised the manuscript. ZN and SI contributed to the development of the study design, interpreted the data, reviewed, edited the manuscript. SMAI contributed to the development of the study design, interpreted the data, reviewed, edited the manuscript. MAB provided intellectual input into the study design, data interpretation, and edited the revised manuscript. MRI provided mental health expertise, contributed to the study idea and design, reviewed the manuscript for important intellectual content, and supervised the whole work. All the authors approved the final version of the manuscript for this submission.

